# What cost decisiveness? A cost benefit analysis of the lockdown to contain COVID-19 in India

**DOI:** 10.1101/2020.07.07.20148338

**Authors:** Mousumi Dutta, Zakir Husain

**Affiliations:** Economics Department, Presidency University

**Keywords:** Cost-benefit analysis, COVID, Interrupted Time Series model, lockdown, SIR model, India

## Abstract

On 24^th^ March, 2020 the Government of India announced a national level lockdown to contain the spread of COVID. The lockdown policy has generated considerable controversy, with critics arguing that it was done without adequate notice or planning, exposed vulnerable section of the population to a humanitarian crisis, and failed to contain the spread of COVID. In response, the Government has claimed that lockdown slowed the transmission process of COVID, thereby reducing the number of cases and deaths substantially. The consequent pressure on the health infrastructure was also much less. To judge between competing claims, this study has undertaken the first cost-benefit analysis of the world’s biggest lockdown. Although the data for a proper cost-benefit analysis is currently not available, we have made a ball point estimate of the net benefit of the lockdown under alternative scenarios. Our estimates reveal the net benefits of lockdown to be negative; moreover, the results are robust under all scenarios.

## 1 Introduction

Following the rapid spread of COVID in Europe and North America in February and early March, 2020, the Government of India (GoI) took prompt and decisive action to contain the spread of COVID by instituting a national lockdown. The lockdown was initially announced for 21 days, but was extended in April and May. A phased withdrawal has been started from June, but restrictions continue in many regions of India.

It is widely accepted that lockdown is a drastic public health measure (Habibi et al. 2020, Sands 2016) with far-reaching consequences (Stone 2020, Polletto et al. 2014). The adoption of such an extreme step requires careful analysis. A cost-benefit analysis of the lockdown in India is thus imperative. While data for a comprehensive cost-benefit analysis will be available only in the coming years, we have made a ball point estimate of the net benefit of the lockdown.

## 2 Contextualizing the lockdown

The lockdown of a society of more than 1.3 billion people over more than two months was hailed by the World Health Organization (WHO) as “tough and timely” (PTI 2020). But the WHO failed to realize the lack of planning associated with the decision. While lockdown is primarily a defensive strategy to buy time for preparing the health infrastructure for the protracted struggle of combating COVID in the post lockdown period, the GoI used it as an offensive weapon— hoping that COVID would peter out during the lockdown. Nor did the GoI appreciate the challenges that would be posed by India’s huge population distributed across diverse geophysical regions, health inequalities, socio-economic disparities, and varied cultural values. The announcement of the lockdown at hours’ notice did not give the huge population, particularly the socio-economically vulnerable section, time to prepare for the coming hardship. It led to a humanitarian crisis, with migrant workers starting a long and disastrous march, with those remaining behind starving, and becoming stricken by COVID. Simultaneously, there was loss of income and livelihoods of millions of people. The education system and maternal and child health programmes, too, were in complete disarray.

The GoI was quick to claim that the lockdown had reduced R_0_, flattened the COVID curve, and prevented the health infrastructure from being over-burdened:

> “The lockdown decision was a timely decision, it helped us contain the positive cases to around 23,000 as of today, while it could have gone up to 73,000. Curve has begun to flatten. The nation has shown that lockdown has been effective in saving lives, containing the infection and slowing down the doubling rate” (Dr. V.K. Paul, cited in *Hindusthan Times*).^1^

A study by Husain *et al*. (2020), however, estimates that hospitalization cases averted by lockdown are only 0.08 to 1.39 lakhs, and would have imposed a burden only in a few states.

Interrupted Time Series models (Wagner et al. 2002) are a popular method for evaluating impact of public health intervention using community level data.^2^ Application of such models to analyse the relationship between lockdowns and trend in COVID cases over time reveal that the first two lockdowns failed to reduce the intercept and slope (Fig. 1); in fact there was a statistically significant increase in both level and slope of the COVID-time curve. Only the third lockdown produced a change in level, though the trajectory of cumulative cases remained unaltered. The fourth lockdown was associated with an increase in both level and trajectory. The lockdowns, therefore, do not seem to have any major impact on the dynamics of COVID transmission. It is possible, however, that COVID would have spread at a greater pace in the absence of the lockdowns; further, there are other implications (both on the cost and benefit side) of lockdowns. The present study incorporates other possible benefits and costs of the strategy to estimate the *net benefit* of the world’s greatest lockdown.

**Figure 1:**
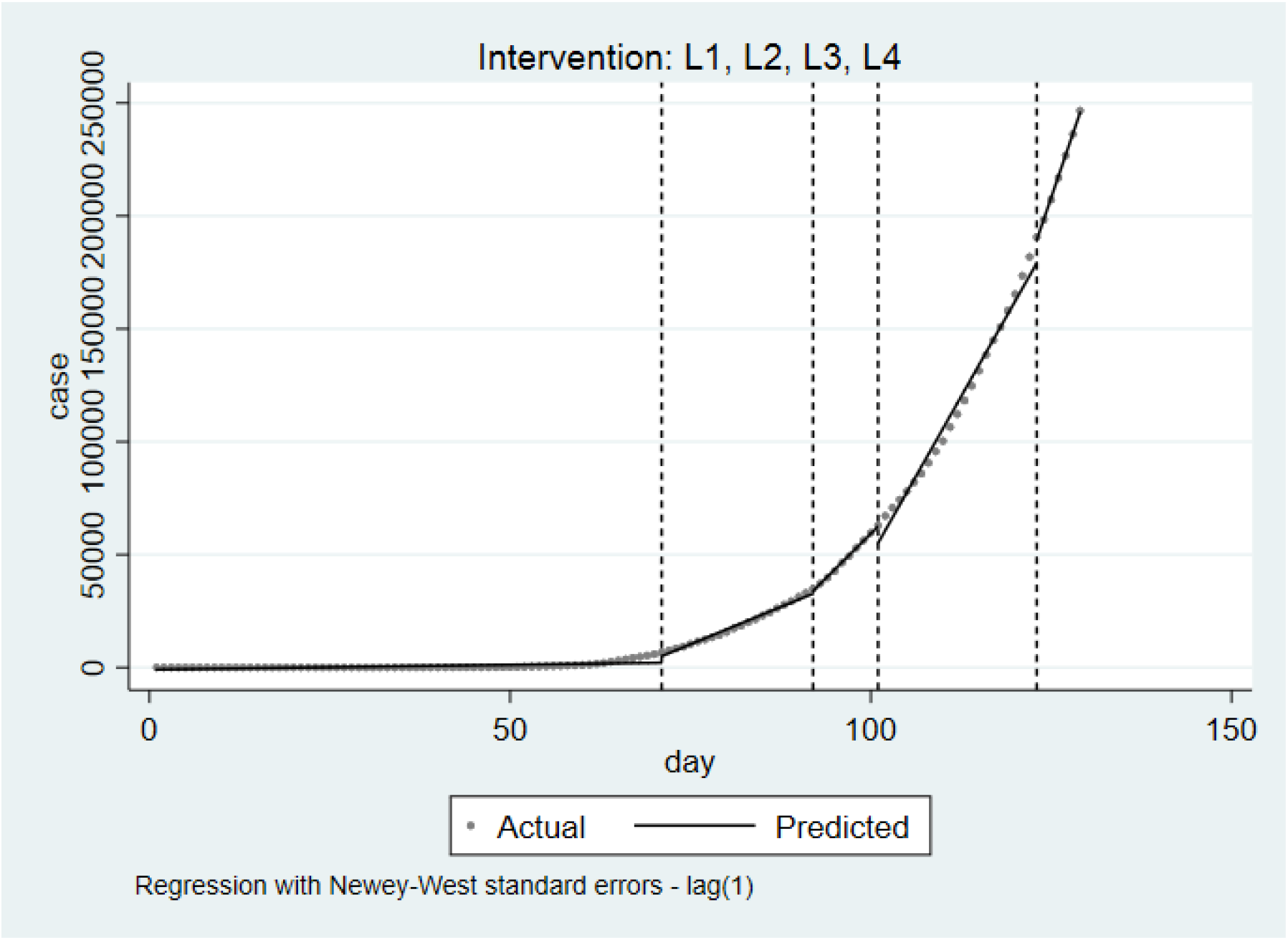
Impact of lockdown on COVID-cases using Interrupted Time Series model.

## 3 Methodology and data

The costs and benefits imposed by the lockdown are categorized in Table 1.

**Table 1:** Costs and benefits of lockdown included in study.

| Benefits | Costs |
| --- | --- |
| 1. COVID cases avoided | 1. Unemployment |
| 2. Net deaths averted due to lockdown:<br>(COVID-deaths + Deaths due to road accidents + Deaths due to Pollution) less (Deaths due to delay in treating cancer and TB + Deaths due to non-COVID reasons<sup>3</sup>) | 2. Loss in production |

Of course, there are other costs of lockdown—increase in domestic violence, disruption of maternal and child health programmes, changes in dietary practices, interruption of education, etc. We have not included such components because there are difficulties in estimating the money value of such costs. It implies that our study underestimates the true cost of lockdown.

The COVID cases averted due to the lockdown is the difference between actual cases as on 6^th^ June 2020, and predictions made using the Susceptible-Infected-Recovered (SIR) model (Kermack and McKendrick 1927). The model parameters are estimated based on Indian data for the pre-lockdown period using a Maximum Likelihood model (R_0_ = 3.048). The costs of treating COVID cases is taken to be Rs.150,000 over a 14 day period (Sethi 2020). The case fatality rate (0.0261%) is taken as the mean of 7-day moving average of deaths per cases, and used to estimate deaths without lockdown.

The shutdown and consequent change in life styles has also changed normal mortality patterns and rates. For instance, reduction of production has led to a cleaner environment, lowering deaths due to pollution. Similarly, reduced vehicular movement has cut down traffic accidents. On the cost side, delays in seeking treatment or disruption of public health programmes is increasing mortality due to cancer, TB, etc. Lockdown is also increasing mental strain, causing suicide rates to shoot up—referred to as “deaths of despair” (Case and Deaton 2020). These have to be incorporated into our analysis.

The decrease in mortality if the reduced pollution level continues for 3 months is estimated to be 0.16 million (Sharma et al. 2020). Based on estimates reported in various studies, we assume an additional 0.15 million TB-related deaths (Stop TB Partnership, Imperial College, Avenir Health, Johns Hopkins University and USAID 2020) and 742 non-COVID related deaths^4^ occurred due to lockdown. Accident deaths averted is estimated by calculating the total number of deaths (mid-year population: 1.38 billion; UNDP estimate of death rate of 7.6564 per thousand), and assuming that 2.9% of them (ICMR, PHFI, and IHME 2017) would have been due to road accidents. Normally 0.22 million deaths would have transpired from cancer deaths;^5^ the *additional* mortality attributable to lockdown (43,849 deaths) is 20% above the normal mortality rate from cancer (Lai et al. 2020).

Combining the mortality-related figures, we get net deaths averted by lockdown to be 285,473. The money value of this is estimated by assuming that such persons would have earned for the next 20 years.^6^ Starting with an initial annual income of Rs. 135,050 (estimated Net National Income per capita for 2019-20 reported by GoI, 2020a), we have assumed that every year there will be an exponential increase of 6, 7 or 8%; a discount rate of 4% is applied to all future payments to obtain present value. These figures, combined with money value of cases avoided, give us three estimates of benefits from lockdown.

We now consider the costs of lockdown. CMIE has estimated that one out of every four worker lost their jobs following lockdown; it implies that about 114 million jobs were lost in April (Vyas 2020), of which 21 million were regained in May (Mishra 2020). Although experts feel that some of these jobs will be lost for a long period, we make the heroic assumptions that 50% of the remaining unemployed will regain work in 2020-21, a further 25% will be re-employed in the next financial year, and job recovery will be complete by April 2022. The money value of unemployment is calculated as the income lost during the jobless period.

The Ministry of Commerce & Industry reports that the core sector has shrunk by 38% due to the lockdown (GoI 2020b). We estimate the production loss under three assumptions—2, 5, and 10% of the Net National Income of Rs. 168.37 trillion (2018-19).

## 4 Was lockdown justified?

The summary of costs and benefits are given in Table 2.

**Table 2:** Summary of costs and benefits of lockdown.

| **Benefit items (Rs. billion)** | **Growth in**<br>**income: 6%** | **Growth in**<br>**income: 7%** | **Growth in**<br>**income: 8%** |
| --- | --- | --- | --- |
| COVID cases avoided | 237.60 | 237.60 | 237.60 |
| Net reduction in mortality | 1132.87 | 1233.91 | 1348.43 |
| **Total Benefit of lockdown** | **1370.47** | **1471.50** | **1586.03** |
| **Cost items (Rs. billion)** | **Loss in**<br>**production: 2%** | **Loss in**<br>**production: 5%** | **Loss in**<br>**production: 10%** |
| Loss due to unemployment | 7199.853 | 7199.853 | 7199.853 |
| Production loss | 841.861 | 2104.652 | 4209.305 |
| **Total Cost of lockdown** | **8041.714** | **9304.506** | **11409.16** |

Net benefits are negative, and vary from Rs.(-) 6.46 trillion to Rs.(-) 10.04 trillion, depending upon the scenario. Even under heroic assumptions, therefore, ball point estimates do not justify the lockdown as costs of the lockdown exceed benefits; moreover, the result holds under *all* the scenarios considered (Table 3).

**Table 3:** Net benefit of lockdown in India (Rs. billion)

| Scenarios |  | Loss in production |  |  |
| --- | --- | --- | --- | --- |
|  |  | 2% | 5% | 10% |
| Growth in income: | 6% | -6,671.25 | -7,934.04 | -10,038.69 |
|  | 7% | -6,570.21 | -7,833.00 | -9,937.66 |
|  | 8% | -6,455.69 | -7,718.48 | -9,823.13 |

## 5 Planning the lockdown

Tyler Cowan had commented that the lockdown was being used by political leaders to shore up their image as strong and decisive decision makers.^7^ In India, in the haste to appear tough and decisive, the GoI failed to plan for the lockdown adequately. Ultimately—with diminishing returns to lockdowns, rising economic costs, and apathy of the public to lockdowns—the GoI had little choice but to start phasing out the lockdown *before* the peak was reached. This policy generates the threat of a major spike in COVID cases.

An alternative policy could have relied on better checking of international passengers at airports, coupled with earlier closing of political boundaries; it might have delayed the entrance of COVID in India. The lockdown itself could have been deferred to May. The postponement would have resulted in more cases—but our projections using the SIR model indicates that the increase in COVID cases would have been a relatively modest 10%. In the interim period, social distancing, and a targeted lockdown in selective hotspots could have geographically contained transmission, while the humanitarian crisis avoided by transporting the migrant workers home. The latter policy had the additional advantage that migrant workers would have been able to return home when few were affected by COVID. It could have possibly avoided the rapid spreading of COVID from states like Maharashtra, Gujarat and Delhi to the districts of Bihar, West Bengal and Jharkhand observed after the *Shramik* trains. In a bid to upstage the states, however, the GoI failed to differentiate between dramatics and achievements; it remains to be seen how costly this is for India.

## Data Availability

Data on COVID-19 cases available from https://api.covid19india.org/csv/. Sources of other data given in paper.

## END NOTES

1 Reported in Hindusthan Times (2020) “Govt lists 3 decisions that changed coronavirus’ course in India”, 24 April. Accessed from https://bit.ly/2Mx4ugp on 6 June.

2 In its simplest form, the ITS model can be presented as: Y_t_ = β_0_ +β_1_T +β_2_X_t_ +β_3_TX_t_ + ε_t_ where Y_t_: the aggregated outcome at time t, in our case the number of new cases of COVID19, T: the time since the start of the study, X_t_: a dummy variable indicating pre-intervention period (coded as 0) or post intervention period (coded as 1), β_0_ represents the intercept or the initial value of the outcome at T=0, β_1_ is the slope or trajectory represents the change in outcome associated with an increase in the time unit (representing the underlying pre-intervention change), β_2_ represents the level change following the intervention, β_3_ indicates the slope change following the intervention (using the interaction between time and intervention), and ε_t_ is the random error term, following a first order autoregressive process.

3 These deaths due to starvation and financial distress, exhaustion, accidents during migration, lack or denial of medical care, suicides, police brutality, crimes, and alcohol-withdrawal.

4 Accessed from https://bit.ly/3eZsUvg on 6 June.

5 It is estimated as 8.3% (ICMR, PHFI and IHME 2017) of estimated deaths for three months.

6 In case of cancer patients, we assume that they would have earned for one year, while TB patients are assumed to earn for 10 years.

7 “The future social and political implications of COVID-19”, Webinar on 10 April, 2020, at Bendheim Center of Finance, Princeton University.

